# “Non-Invasive Imaging Modalities in Coronary Artery Disease: *A meta analysis* comparing *CCTA* and *Standard of Care (SOC)”*

**DOI:** 10.1101/2023.06.14.23291404

**Authors:** Avichal Dani, Pari Shah, Dev Desai

## Abstract

**Introduction:** Coronary Artery Disease has become a global pandemic and a major cause of death especially among Older population. The risk factor calculation or the degree of Coronary artery damage is usually an invasive procedure. A paradigm change has occurred in the last ten years regarding the assessment of coronary artery disease (CAD) utilising coronary computed tomography angiography (CCTA).

**Aims and Objectives:** To Comapre CCTA with Standard Care to calculate the need of revascularization, invasive coronary angiography as well as to compare both for MI and All cause mortality

**Results:** CCTA was found to be associated with a significant increase in revascularizations (RR =1.401, 95% CI =1.315-1.492, p<0.001) as well as invasive coronary angiography procedures (RR =1.304, 95% CI =1.208-1.409, p<0.001). However, it was associated with decreased incidences of MI (RR =0.752, 95% CI =0.578-1.409, p<0.033). There was no significant association with all-cause mortality.

**Conclusion:** CCTA is significantly correlated with a reduction in MI episodes and an increase in revascularizations and ICA procedures. However, it was found that it had no effect on all-cause mortality. On the contrary, standard care approaches were associated with greater rates of MI but lesser referrals for invasive coronary angiography and revascularization.

## INTRODUCTION

Coronary artery disease is a true epidemic of the modern era, one of the many maladies afflicting the human population. It is recognised as a leading cause of death in both developed and developing countries. Years of life lost (YLLs) and disability-adjusted life years (DALYs) due to CHD have been showing growing trends in India, according to the World Health Organization (WHO) and Global Burden of Disease Study. Studies show that the prevalence of CHD varies between 1% and 2% in rural and 2% to 4% in urban populations. Trends in Coronary Heart Disease Epidemiology in India. [1]

The symptoms of CAD might range from none to chest discomfort, breathlessness, or heart attack. [2]The difficulty of quickly making an accurate diagnosis is exacerbated by the prevalence of unusual symptoms and clinical presentations. As a result, diagnostic methods used in emergency department (ED) chest pain units are based on the standard of care (SOC). [3]The current emergency department (ED) standard care (SC) for risk stratification of ACPS patients in many institutions includes clinical data, serial electrocardiograms (ECGs), and cardiac biomarkers. Although time-consuming, this strategy has minimised diagnostic errors. A paradigm change has occurred in the last ten years regarding the assessment of coronary artery disease (CAD) utilising coronary computed tomography angiography (CCTA).[4] The clinical value of CCTA at different phases of CAD, from the diagnosis of early subclinical illness to the evaluation of acute chest pain, is being supported by growing evidence. Additionally, CCTA can be utilised for non-invasive plaque burden quantification and high-risk plaque identification, which can help with diagnosis, prognosis, and treatment. With the aid of cutting-edge technologies derived from CCTA, atherosclerotic plaque growth may be understood, and patients with CAD can benefit from risk classification and quick medical decision-making. Advancements have allowed for minimal radiation exposure, effective coronary characterization, and detailed imaging of atherosclerosis over time. Thus, CCTA offers a focal point for a multidisciplinary approach that includes immunology, pathology, radiology, and cardiology to deepen our understanding of CAD and enhance patient management.

The significance of CCTA as a non-invasive diagnostic technique in improving diagnoses and changing treatments and investigations is becoming more and more explicit, however, it is still ambiguous whether this change in CAD management over the standard of care is feasible.[5] The impact of CCTA on clinical care and the usage of downstream resources must therefore be addressed.

## METHODOLOGY

### Data source

The following databases were searched electronically: PubMed, EMBASE, OVID, Web of Science, Cochrane Library, Google Scholar, and Controlled Trials Meta Register. “Coronary artery disease”, “functional stress testing”, “angiography”, and “CCTA vs SOC” were text keywords. Manually scanning the reference lists of applicable retrieved publications led to the discovery of more investigations. English was the sole permitted language. Human investigations were the only ones that yielded results.

### Eligibility Criteria

The researchers looked for randomized controlled studies that compared the Coronary Computed Tomography (CCTA) and Standard of Care (SOC) approach in the management of CAD and the associated outcomes. Research papers were also excluded from the analysis if it was impossible to extract the appropriate data from the published articles; there was significant overlap between authors, institutes, or patients in the published literature; the measured outcomes were not clearly presented in the literature; the measured outcomes were not clear or articles were written in a language other than English. These exclusion criteria also applied to letters, comments, editorials, expert opinions, reviews without original data, and case reports.

### Study Identification

The author examined each title and abstract that the search strategy found. Two non-author independent reviewers also evaluated them for adherence to the eligibility requirements.

### Data extraction

Each qualified manuscript was reviewed by two independent reviewers separately. The number of patients, their age, gender, type of intervention, and incidence of revascularisation, invasive coronary angiography procedures, all-cause mortality and myocardial infarction were all collected from each manuscript. Conflicts were addressed by further discussion or consultation with the author and a third party. The modified Jadad score was used to measure the study’s quality.

### Outcome Measures

Our primary outcomes were the incidence of revascularisation, invasive coronary angiography procedures, all-cause mortality, and myocardial infarction.

### Statistical analysis

All of the information was gathered and placed into software for analysis. RevMan 5.3 was used for appropriate statistical tests. To analyse important clinical outcomes, fixed- or random-effects models were used to generate a mean difference, standardized mean difference (SMD), odds ratios, and relative risk (RR) with 9per centnt confidence intervals (CIs). Statistical heterogeneity was measured with the χ^2^; P < 0.100 was considered as a representation of significant difference. I^2^ greater than or equal to 50% indicated the presence of heterogeneity. A statistically significant difference was defined as P<0.05.

**chart 1:**
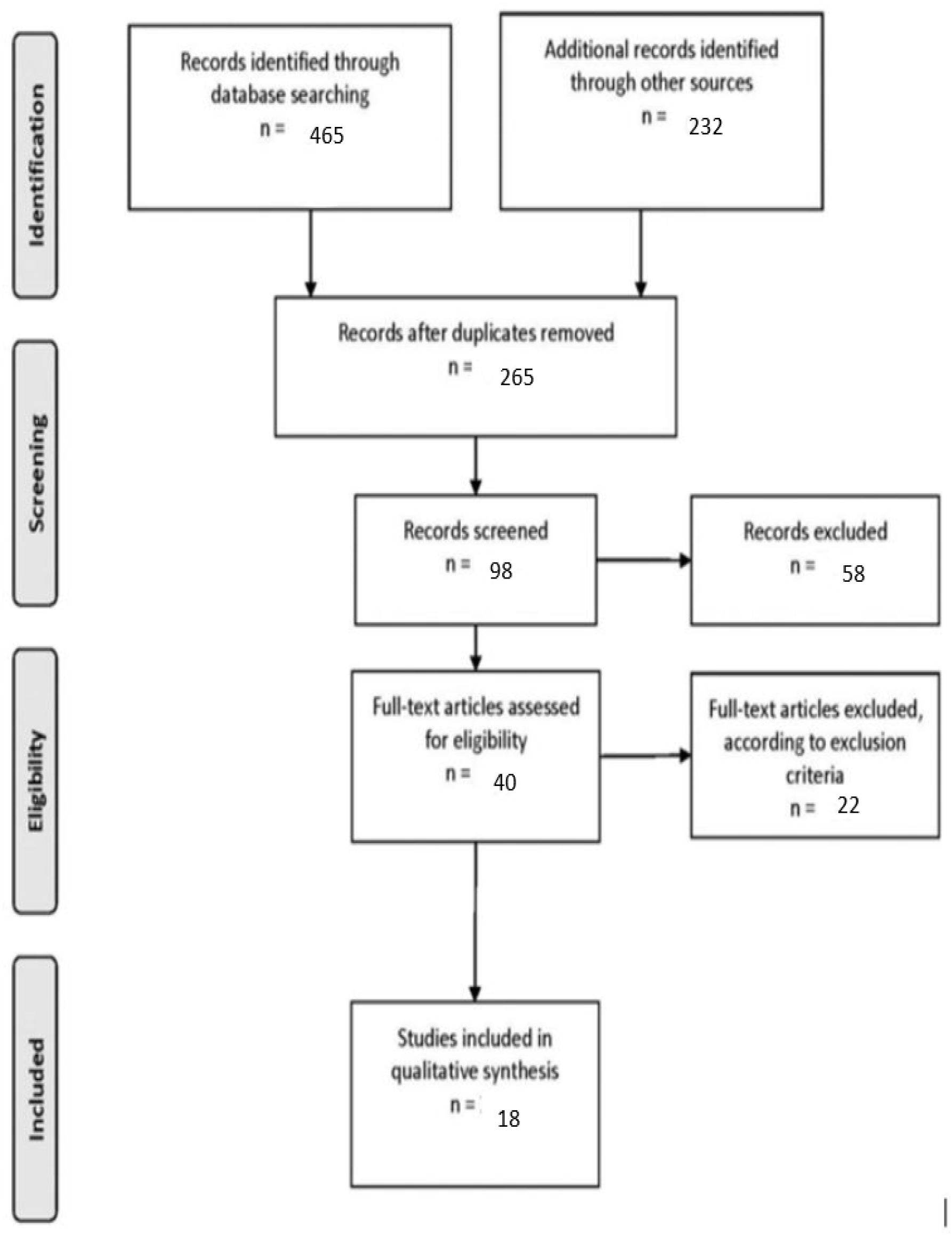
PRISMA flow chart

## RESULTS

As seen in Figure 1 and 4 respectively, coronary computed tomography angiography (CCTA) was found to be associated with a significant increase in revascularizations (RR =1.401, 95% CI =1.315-1.492, p<0.001) as well as invasive coronary angiography procedures (RR =1.304, 95% CI =1.208-1.409, p<0.001). However, as per Figure 3, it was associated with decreased incidences of MI (RR =0.752, 95% CI =0.578-1.409, p<0.033). There was no significant association with all-cause mortality (Figure 2).

**Figure 1:**
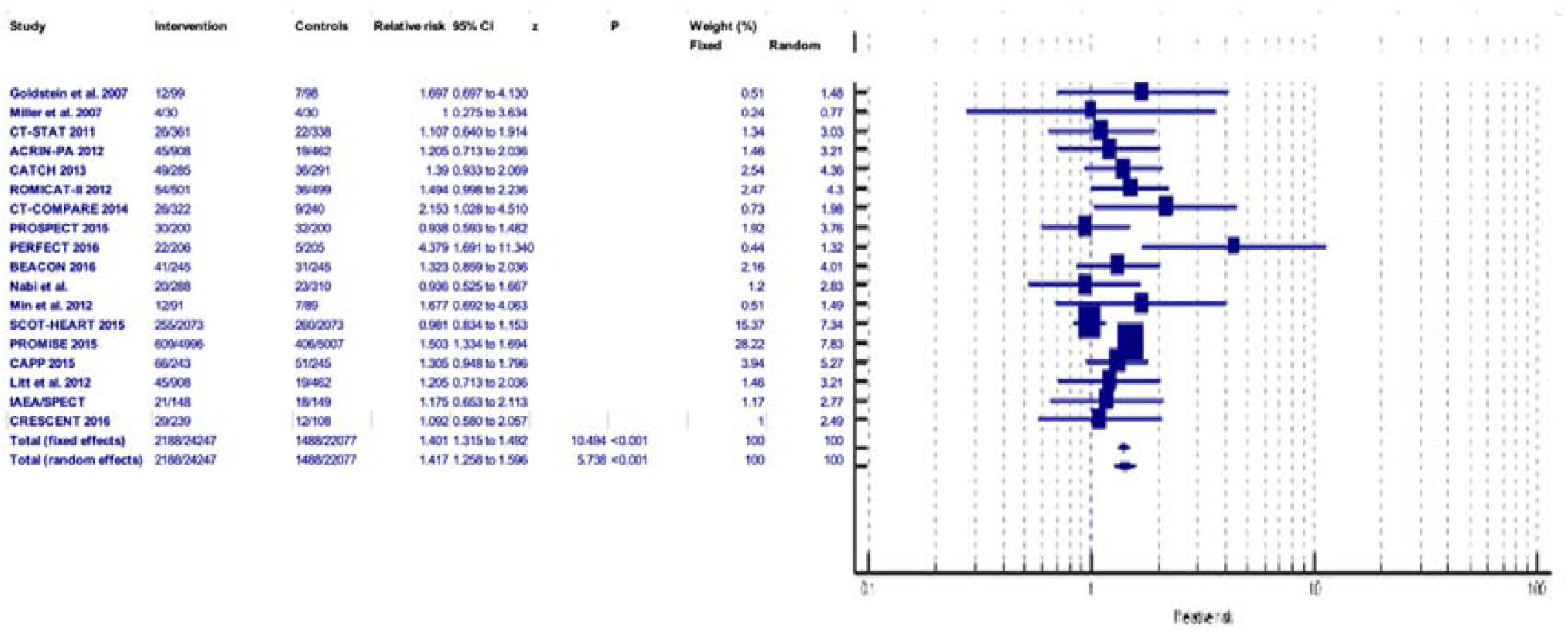
-REVAS

**Figure 2:**
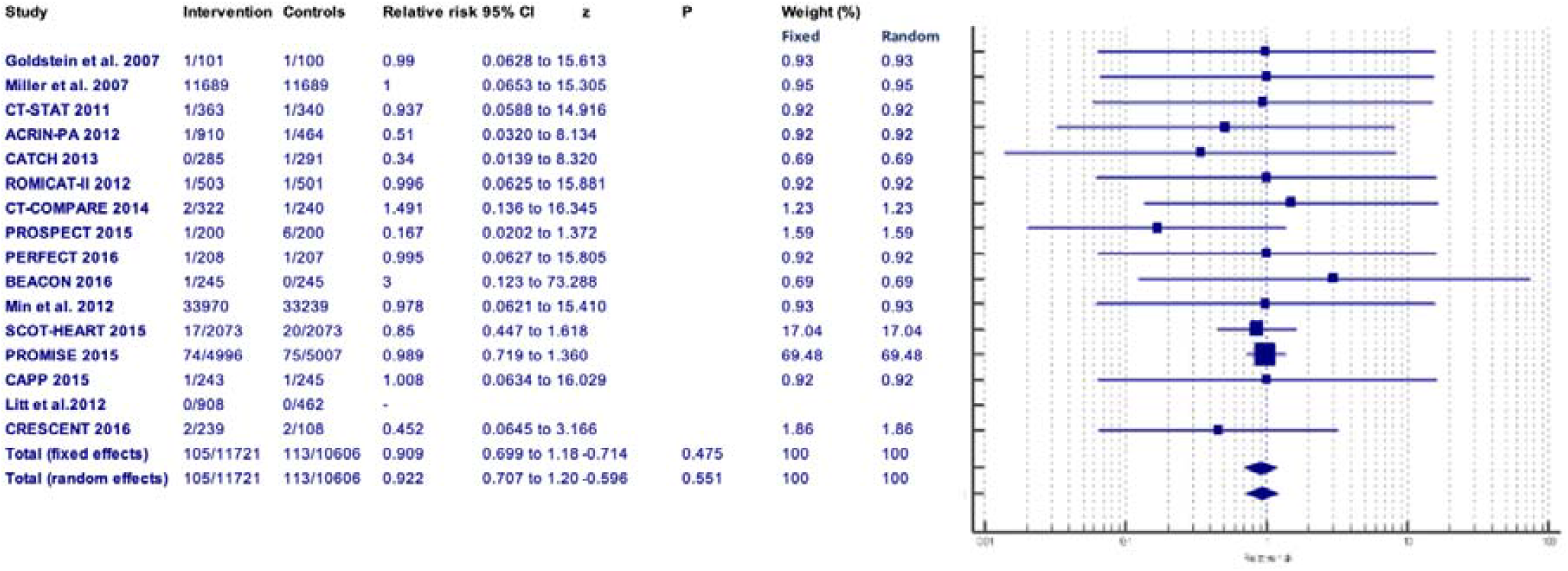
-ACM

**Figure 3:**
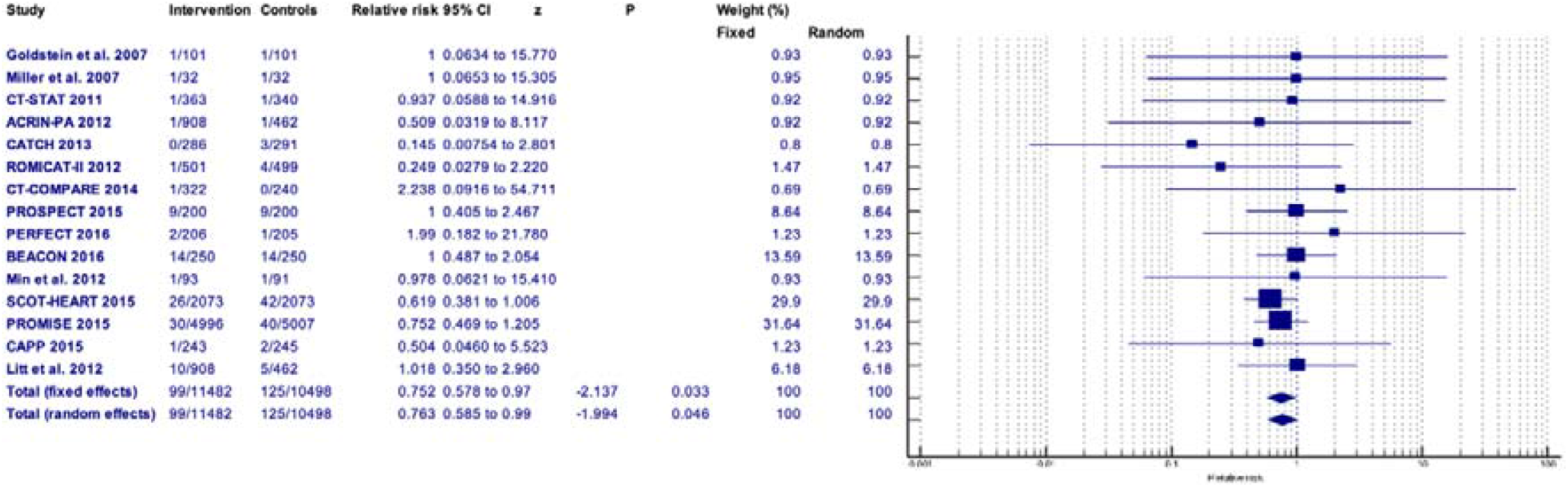
-MI

**Figure 4:**
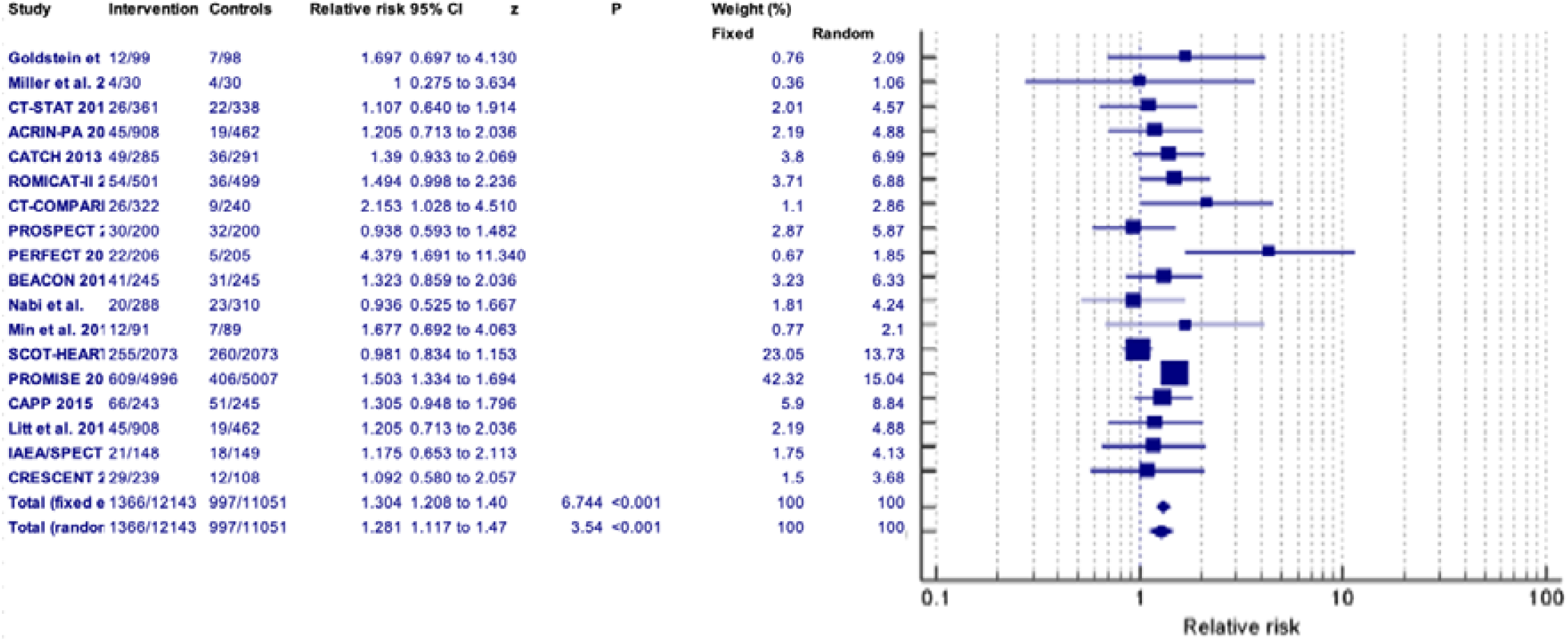
-ICA

Comparison 1:-Revascularisation

Comparison 2:-All cause mortality

Comparison 3:-Myocardial Infarction

Comparison 4:-Invasive coronary angiography

## DISCUSSION

Coronary computed tomography angiography (CCTA) is a non-invasive alternative tool for the diagnosis of obstructive CAD.[5]This tool has been evaluated in randomized controlled trials (RCTs) in patients with stable chest pain and an intermediate probability of CAD. However, there is uncertainty regarding its usefulness in reducing cardiovascular outcomes in the patient population.[6]

Prior studies of patients with stable chest pain undergoing diagnostic imaging have primarily focused on diagnostic and prognostic performance, with relative neglect of patient-centred outcomes. [7]

As it can be seen in Figure 1, we found CCTA to be associated with higher incidences of revascularisation procedures than the standard care approach, which is supported by many studies[6], [8], [9] and [10]. However, some studies [11], [12]did report no difference in revascularisation incidence between the two groups.

As per Figure 2, there is no difference in all-cause mortality statistics between the two groups similar to the results of other studies.[9],[12],[10]

One can infer from Figure 3 that CCTA was associated with decreased incidences of myocardial infarction which is an event that is clearly undesirable. This is in agreement with the results of one paper [10] but does not conform to the findings of another [12], which reported no difference between the two groups.

Figure 4 shows increased rates of invasive coronary angiography procedures in the CCTA arm, which follows the findings of various papers[6], [9], [10] and [12] We did find one study [8] that showed no notable difference.

Altogether we found more papers with results in line with ours and only a few with contrasting results.

It goes without saying that lowered MI incidence associated with CCTA is a tremendously positive outcome. What still remains unknown is whether the rise in invasive treatments (revascularisation and ICA) improves patient outcomes or lessens the need for future additional testing.

## CONCLUSION

CCTA is significantly correlated with a reduction in MI episodes and an increase in revascularizations and ICA procedures. However, it was found that it had no effect on all-cause mortality. On the contrary, standard care approaches were associated with greater rates of MI but lesser referrals for invasive coronary angiography and revascularization. The choice of the modality should be on the judgement of the attending physician and should be dependant on the condition of each patient as well as other criterias and morbidities should be a part of the decision.

**Table 1:** -Description of studies

| No . | Study | Type | Country | Publish year | Study period | Sample size | Intervention | Follow up | Q u a l i t y A s s e s s m e n t |
| --- | --- | --- | --- | --- | --- | --- | --- | --- | --- |
| 1 | **Goldstein et al. 2007 [13]** | RCT | USA | 2007 | March to September 2005 | 203 | Multi-Slice Computed Tomography (MSCT) or Standard of Care (SOC) | 6 months | G o o d |
| 2 | **Miller et al. 2007 [14]** | RCT | USA | 2007 | 2008-2010 | 76 | FAC/Nylon | 3-4 mo | G o o d |
| 3 | **CT-STAT 2011** [15] | RCT | USA | 2011 | June 2007 and November 2008 | 699 | CCTA or myocardial perfusion imaging (MPI) | 6 months | G<br>o<br>o<br>d |
| 4 | **ACRIN-PA 2012**[16] | RCT | USA | 2012 | NA | 1370 | CCTA or traditional care | 1 month | G<br>o<br>o<br>d |
| 5 | **CATCH 2013** [17] | RCT | Denmark | 2013 | January 2010 to January 2013, | 600 | CCTA or standard care | 4 months | G<br>o<br>o<br>d |
| 6 | **ROMIC AT-II 2012** [18] | RCT | USA | 2012 | April 2010 – March 2012 | 1000 | CCTA or standard evaluation | 28 days | G<br>o<br>o<br>d |
| 7 | **CT-COMPA RE 2014** [19] | RCT | Australia | 2014 | March 2010 to April 2011 | 562 | CCTA or exercise test ECG | 1 month | G<br>o<br>o<br>d |
| 8 | **PROSPECT 2015** [20] | RCT | USA | 2015 | July 2008 to March 2012 | 400 | CCTA or radionuclide stress myocardial perfusion imaging (MPI). | 1 year | G<br>o<br>o<br>d |
| 9 | **PERFECT 2016** [21] | RCT | USA | 2017 | July 2011 and December 2013 | 411 | CCTA or stress testing | 1 year | G<br>o<br>o<br>d |
| 10 | **BEACON 2016** [22] | RCT | Netherlands | 2016 | July 2011 to February 2015 | 500 | CCTA and standard care | 1 month and 6 months | G<br>o<br>o<br>d |
| 11 | **Nabi et al. 2016** [23] | RCT | USA | 2016 | NA | 598 | Stress gated tomographic myocardial perfusion imaging (SPECT) or CCTA | 6.5 months | Good |
| 12 | **Min et al. 2012** [7] | RCT | USA | 2012 | December 2008 and June 2009 | 180 | CCTA or myocardial perfusion imaging | 2 months | Good |
| 13 | **SCOT-HEART 2015** [24] | RCT | Scotland | 2015 | Nov 18, 2010, and Sept 24, 2014 | 4146 | standard care plus CCTA or standard care alone | 6 weeks | Good |
| 14 | **PROMISE 2015** [25] | RCT | USA | 2015 | NA | 10,003 | CCTA or ECG or Stress echocardiography or Nuclear stress testing | Median follow-up was 25 months with a maximum follow-up of 50 months. | Good |
| 15 | **CAPP 2015** [26] | RCT | Ireland | 2015 | September 2010 to November 2011 | 500 | Exercise stress electrocardiogram test or CCTA | 3 months and 1 year | Good |
| 16 | **Litt et al. 2012** [27], [28] | RCT | USA | 2012 | NA | 1370 | CCTA or traditional care | 1 month | Good |
| 17 | **IAEA/SPECT-CTA 2017** [28] | RCT | Brazil, Czech Republic, India, Mexico, Slovenia, and Turkey | 2017 | June 2011 and 2014 | 303 | myocardial perfusion imaging (MPI) or CCTA | 6 months | Good |
| 18 | **CRESCE**<br>**NT 2016**<br>[29] | RCT | Netherla<br>nds | 2016 | April 2011<br>and July<br>2013 | 350 | Cardiac CT<br>or<br>Functional<br>testing | 1<br>year | G<br>o<br>o<br>d |

## Data Availability

All data produced in the present work are contained in the manuscript

